# Impact of Dexmedetomidine on the Prognosis of Patients with Sepsis-Induced Myocardial Injury: A Retrospective Cohort Study

**DOI:** 10.1101/2025.02.11.25321833

**Authors:** Zhang Xinyi, Guo Xiaoyan, Zhao Zhongxing, Li Wei, Chen Huaiyu, Li Hongyan, Lin Jiandong, Lin Mingrui

## Abstract

**Background:** Sepsis-Induced Myocardial Injury (SIMI) is a serious complication with high in-hospital and 28-day mortality. Dexmedetomidine (DEX), a selective α2-adrenergic receptor agonist, has shown anti-inflammatory and cardioprotective effects, but its impact on SIMI prognosis is unclear.

**Methods:** This retrospective study used the MIMIC-IV database, including SIMI patients diagnosed per Sepsis-3 criteria. Propensity score matching (PSM) balanced baseline characteristics, and Cox regression analyzed the association between DEX use and mortality outcomes. External validation was performed using the MIMIC-III database.

**Results:** Among 3,921 SIMI patients (375 DEX, 3,546 non-DEX), DEX use showed a trend toward lower 28-day mortality (HR 0.49, 95% CI 0.23–1.05, p = 0.068) and in-hospital mortality (HR 0.45, 95% CI 0.20–1.02, p = 0.055). After PSM, mortality remained lower in the DEX group. ICU and hospital stays were longer in the DEX group, but CRRT use showed no significant differences. External validation confirmed these findings.

**Conclusion:** DEX use may reduce 28-day and in-hospital mortality in SIMI patients, providing preliminary evidence for its potential benefits. Further large-scale studies are needed to confirm these results.

## Introduction

Sepsis is a life-threatening condition defined by multiple-organ dysfunction resulting from an excessive host response to infection. According to the Global Burden of Disease Study, sepsis impacts over 49 million people annually and is responsible for 19.7% of deaths worldwide^1^. Despite recent improvements in the prognosis of septic patients, the mortality rate remains alarmingly high, ranging from 40% to 50%^2^. Both clinical and basic research have demonstrated that sepsis significantly impacts the cardiovascular system^3^. Cardiac dysfunction, with sepsis-induced myocardial injury (SIMI) being a prominent example, is regarded as a critical determinant of the high mortality associated with sepsis^4^. SIMI, though recognized as a potentially reversible complication during sepsis progression, is associated with a poor prognosis. Patients face an in-hospital mortality rate of 35% and a 1-year mortality rate as high as 51%^5^. Given the high mortality rates, effective management strategies are urgently needed to improve outcomes.At present, the management of SIMI mainly emphasizes supportive care. Recent research has highlighted the application of positive inotropic agents (e.g., dopamine and levosimendan), beta-blockers, and anti-inflammatory therapies in the treatment of SIMI^6–8^.

Dexmedetomidine (DEX), a highly selective α2-adrenergic receptor agonist, is commonly utilized for analgesia and sedation in ICU patients. In recent years, numerous studies have explored the potential benefits and drawbacks of DEX in various clinical settings, particularly its effects on cardiac function and myocardial injury.The impact of DEX on myocardial injury remains a subject of debate. Tianyi et al. ^9^ demonstrated that DEX mitigates septic myocardial injury by modulating macrophage-derived exosomal miR-29b-3p and enhancing autophagy in cardiomyocytes via the regulation of glycogen synthase kinase 3β. Additionally, a clinical study confirmed that DEX can reduce heart rate and inflammatory markers, protect cardiomyocytes, and improve cardiac function in patients with sepsis and myocardial injury^10^. Despite its reported benefits, other studies have raised concerns regarding its safety and effectiveness in specific contexts.The study by Liaoyuan et al. demonstrated that compared to other sedatives (propofol or midazolam), DEX increases mortality and exacerbates myocardial dysfunction in septic patients, potentially through promoting inflammatory responses by directly acting on the heart^11^. Another study revealed that DEX exacerbates metabolic disturbances in septic cardiomyopathy through an α2A-adrenoceptor-dependent mechanism, thereby worsening cardiac dysfunction^12^. Therefore, further research is essential to clarify the dual effects of DEX on cardiac function and coagulation, focusing on its mechanisms of action and patient-specific risk factor.

Numerous studies have confirmed that DEX provides critical protection against multi-system organ damage in sepsis by modulating inflammation and apoptosis^13–15^. However, evidence on the impact of DEX on the survival outcomes of patients with SIMI remains scarce. Therefore, this study aims to explore the potential association between DEX use and the prognosis of patients with SIMI through a large retrospective dataset.

## Material and methods

### Data sources and Ethics approval

This study is a retrospective observational analysis utilizing the MIMIC-IV database (Information Mart for Intensive Care IV), an open-access dataset that includes hospitalization records of more than 65,000 ICU patients from 2008 to 2022. Version 3.1 of the MIMIC-IV database, released on October 11, 2024, offers several improvements over the previous MIMIC-III version^16,17^. These enhancements encompass comprehensive updates to laboratory test results, detailed medication records (e.g., NSAID and ACEI usage), as well as more precise documentation of vital signs and patient scoring systems.In compliance with the Health Insurance Portability and Accountability Act (HIPAA), all identifiable patient information within the database has been anonymized to safeguard privacy. As a result, this study does not require patient consent or ethical approval. Zhang Xinyi, a member of our research team, was granted access to the database and took responsibility for data extraction (Authorization Code: 10320923).

### Study design and study population

This study is a retrospective observational study. The inclusion criteria were as follows: patients diagnosed with sepsis according to the Sepsis-3 definition (life-threatening organ dysfunction caused by a dysregulated host response to infection with an increase in SOFA score ≥2 from baseline) and aged between 18 and 80 years^18^. The exclusion criteria included the following conditions: the presence of other cardiovascular diseases (such as coronary artery disease, chronic obstructive pulmonary disease, chronic heart failure, valvular heart disease, non-sepsis-related cardiomyopathy, and myocarditis), a history of kidney disease, absence of troponin T test results, and patients with DEX use ≤4 hours or >48 hours. For patients with multiple ICU admissions, only the first hospitalization and the first day’s records were included. For patients with multiple ICU admissions, only the first hospitalization and the first day’s records were included.Retrospective data collection for eligible patients included the following: demographic characteristics (age, weight, sex, race); disease severity (SOFA score and SAPS II score); treatment measures during hospitalization (use of fentanyl, midazolam, propofol, and application of CRRT); laboratory indicators (minimum hemoglobin, minimum platelet count, minimum and maximum white blood cell count, maximum blood urea nitrogen, maximum creatinine, minimum and maximum levels of calcium, sodium, potassium, and glucose); vital signs (mean heart rate, systolic blood pressure, diastolic blood pressure, mean arterial pressure, respiratory rate, body temperature, and oxygen saturation); and outcome events (in-hospital mortality, 28-day mortality, ICU length of stay, total hospital length of stay). Currently, there is no universally accepted definition of SIMI, but studies have confirmed a significant association between troponin T levels and mortality in septic patients^19^, Therefore, this study defines SIMI as a troponin T level ≥0.01 ng/mL within 24 hours^20^.

### Data management and statistical analysis

This study utilized DBeaver software (version 24.1.4.202408041450) and structured query language (SQL) to extract data from the MIMIC-IV 3.1 database. The proportion of missing values for each variable was less than 5%. To ensure data completeness and availability, continuous variables were imputed using the median, while categorical variables were imputed using the mode. Patients with SIMI who received DEX were designated as the experimental group, while those who did not receive DEX formed the control group. Baseline characteristics were compared using the Mann-Whitney U test for continuous variables, with results expressed as medians and interquartile ranges, and the chi-square test for categorical variables. To balance baseline characteristics between the two groups, propensity score matching (PSM) was employed, with a caliper value set at 0.2 for nearest-neighbor matching. Standardized mean difference (SMD) was used to evaluate the balance of baseline characteristics after matching. Kaplan-Meier survival curves were generated to visualize in-hospital and 28-day mortality rates, and differences between groups were assessed using the log-rank test. Cox proportional hazards regression models were used to analyze the association between DEX use and mortality outcomes, with hazard ratios (HR) and 95% confidence intervals (CI) calculated. The analysis included three models: Model 1 was unadjusted, Model 2 was partially adjusted (accounting for age, sex, SOFA score, and SAPS II score), and Model 3 was fully adjusted (including laboratory indicators, vital signs, and treatment measures such as the use of propofol, midazolam, fentanyl, and CRRT). Additionally, subgroup analyses were conducted based on key clinical and demographic characteristics, including age (≤60 years vs. >60 years), sex, SOFA score (≤8 points vs. >8 points), SAPS II score (≤50 points vs. >50 points), and potassium levels (normal/low vs. high). HRs and 95% CIs were calculated for each subgroup. All statistical analyses were performed using R software (version 4.4.2), and a two-sided P-value <0.05 was considered statistically significant.

### External validation

External validation was performed using the MIMIC-III (Medical Information Mart for Intensive Care) database, which is a publicly available critical care database containing de-identified health data from Beth Israel Deaconess Medical Center. MIMIC-III includes records of over 40,000 ICU patients from 2001 to 2012^21^. Our research team member, Zhang Xinyi, obtained access to the database after completing the required training on human subject research protection (Authorization Code: 10320923).

The external validation cohort consisted of 1,282 ICU patients from the MIMIC-III database who were diagnosed with SIMI. To ensure a sufficient number of eligible patients, we applied the same inclusion and exclusion criteria as those used in MIMIC-IV, excluding minors under 18 years old and patients who used dexmedetomidine for more than 48 hours. A Cox proportional hazards regression model was used to assess the relationship between dexmedetomidine use and in-hospital as well as 28-day mortality outcomes.

## Results

### Patient characteristics

During the study period, 16,937 cases of SIMI were identified, as shown in Figure 1. After applying the exclusion criteria, 3,921 patients were deemed eligible for analysis, comprising 375 patients in the DEX group and 3,546 patients in the non-DEX group.

**Figure 1:**
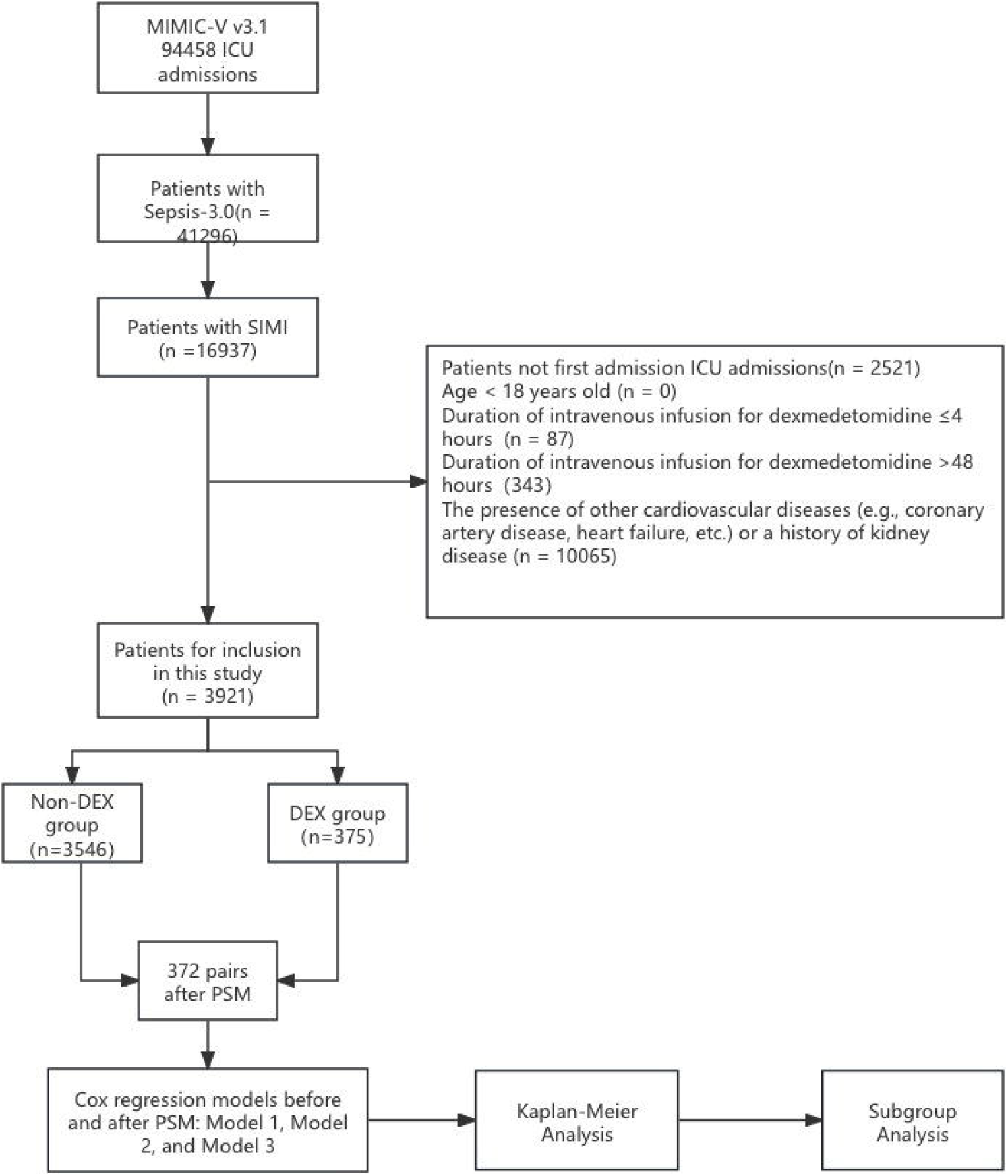
Flowchart of the study Abbreviations: ICU: Intensive Care Unit;MIMIC-V v3.1: Medical Information Mart for Intensive Care IV ;Sepsis-3.0: Sepsis, defined according to the third international consensus definition;PSM: Propensity Score Matching;Non-DEX: Patients not treated with Dexmedetomidine

Significant differences were observed between the DEX and non-DEX groups concerning age, gender, race, and weight (Table 1). The DEX group had higher SOFA scores and showed notably greater usage rates of fentanyl, midazolam, and propofol compared to the non-DEX group. Laboratory findings also revealed that blood urea nitrogen (BUN) levels were significantly lower in the DEX group. However, no significant difference was found in the use of continuous renal replacement therapy (CRRT) between the groups. Following propensity score matching, 372 patients treated with DEX were matched to 372 patients who did not receive it. The matched cohort displayed well-balanced baseline characteristics, with Standardized Mean Differences (SMD) for all variables below 10%.

**TABLE 1:** Baseline Characteristics Between DEX and Non-DEX Groups Before and After Propensity Score Matching. Abbreviations: SMD: Standardized Mean Difference; SBP: Systolic Blood Pressure; DBP: Diastolic Blood Pressure; MBP: Mean Blood Pressure; SpO_2_: Oxygen Saturation; SOFA: Sequential Organ Failure Assessment; SAPSII: Simplified Acute Physiology Score II; WBC: White Blood Cell; BUN: Blood Urea Nitrogen; CRRT: Continuous Renal Replacement Therapy.

Analysis revealed that the DEX group had significantly lower in-hospital and 28-day mortality rates compared to the non-DEX group, irrespective of whether the data was analyzed before or after propensity score matching (PSM). Furthermore, the DEX group experienced notably longer ICU and overall hospital stays, as well as extended 28-day survival times. Prior to PSM, the in-hospital mortality rate in the DEX group was 15.2%, compared to 27.1% in the non-DEX group, while the 28-day mortality rates were 18.1% and 28.6%, respectively. This pattern remained consistent even after PSM (Table 2).

**TABLE 2:** Comparison of Clinical Outcomes Between DEX and Non-DEX Groups Before and After Propensity Score Matching. Abbreviations: ICU: Intensive Care Unit;p: p-value (statistical significance)

### Relationship between outcomes and dexmedetomidine use

Our study utilized Cox regression models to assess the association between DEX administration and both 28-day hospital mortality and in-hospital mortality, including analyses conducted before and after propensity score matching (PSM). Before PSM, the use of DEX was significantly associated with a reduction in 28-day hospital mortality, as indicated by the following results: unadjusted model [HR, 0.57 (0.447–0.731, p = 8.01e-06)], partially adjusted model [HR, 0.53 (0.411–0.674, p = 3.53e-07)], and fully adjusted model [HR, 0.57 (0.443–0.739, p = 1.94e-05)]. Similarly, for in-hospital mortality, the unadjusted model [HR, 0.42 (0.318–0.544, p = 1.33e-10)], partially adjusted model [HR, 0.40 (0.309–0.529, p = 3.81e-11)], and fully adjusted model [HR, 0.44 (0.333–0.579, p = 5.59e-09)] all demonstrated a significant risk reduction.After PSM, the protective effect of DEX remained consistent. For 28-day hospital mortality, the unadjusted model [HR, 0.64 (0.469–0.873, p = 0.00487)], partially adjusted model [HR, 0.65 (0.477–0.889, p = 0.00697)], and fully adjusted model [HR, 0.58 (0.417–0.798, p = 0.00089)] showed significant associations. Likewise, for in-hospital mortality, the unadjusted model [HR, 0.46 (0.329–0.632, p = 2.29e-06)], partially adjusted model [HR, 0.48 (0.345–0.663, p = 9.81e-06)], and fully adjusted model [HR, 0.41 (0.288–0.577, p = 4.13e-07)] consistently indicated a lower risk(Table 3).

**Table 3:** Survival Outcomes of DEX Users and Non-Users in SIMI Patients Before and After Propensity Score Matching. Abbreviations: HR: Hazard Ratio;CI: Confidence Interval ;p-value: Probability Value

Model 1: Unadjusted model, including only the DEX and non-DEX group variables.

Model 2: Adjusted for key demographic variables, including age, gender, and SOFA,SAPSII.

Model 3: Fully adjusted model, accounting for confounding variables with p-value <0.05 in univariate analysis. These variables include age, gender, ethnicity, CRRT, propofol, midazolam, fentanyl, respiratory rate, SpO_2_, BUN, creatinine, SOFA, and others.

After propensity score matching (PSM), Figures 2A and 2B present the Kaplan-Meier curves for in-hospital mortality and 28-day mortality, respectively, indicating that the 28-day survival rate in the DEX group was significantly higher than that in the non-DEX group (HR, 0.64 [0.469– 0.873], log-rank test: p = 0.00487). Additionally, the in-hospital mortality risk in the DEX group was significantly reduced (HR, 0.46 [0.329–0.632], log-rank test: p = 2.29e-06).

**Figure 2:**
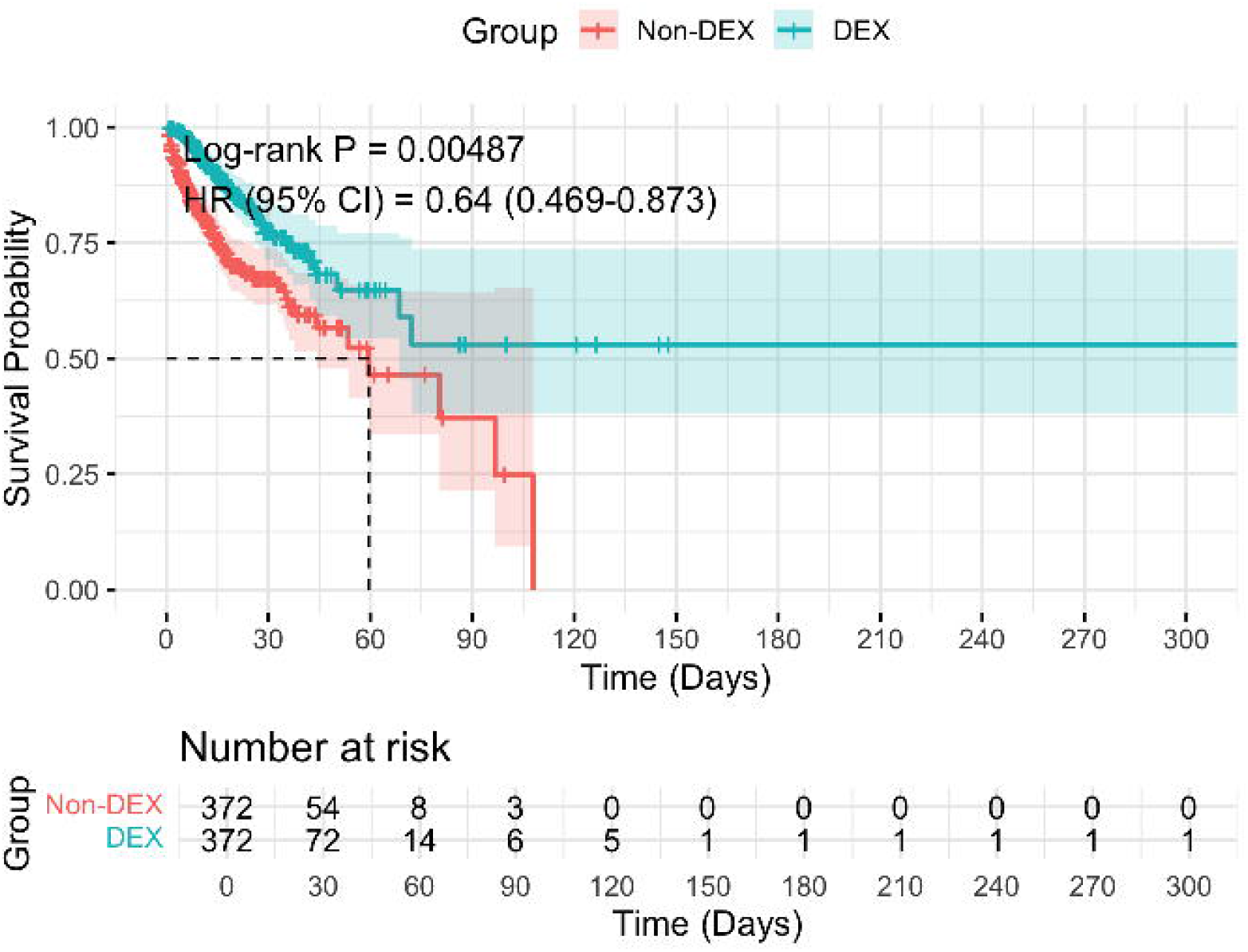
A: Kaplan-Meier Survival Curve for 28-Day Mortality Comparing DEX and Non-DEX Groups After PSM Figure B: Kaplan-Meier Survival Curve for In-Hospital Mortality Comparing DEX and Non-DEX Groups After PSM

The blue line represents DEX users, while the red line corresponds to non-DEX users. To validate the robustness of the study results against potential unmeasured or residual confounding factors, a sensitivity analysis was conducted using E-values to quantify the sensitivity of the results to unmeasured confounders (https://www.evalue-calculator.com/evalue/)^21,22^. The findings revealed that the association between DEX treatment and 28-day hospital mortality is robust, with a hazard ratio (HR) of 0.64. To entirely explain away this observed association, the relative risk between unmeasured or residual confounders and both the exposure and the outcome would need to exceed 2.06 (point estimate) and 1.43 (confidence interval lower limit), respectively. For in-hospital mortality, the E-value analysis showed a point estimate E-value of 2.8 and a confidence interval E-value of 2.09. This indicates that only if the association strength (risk ratio) between unmeasured or residual confounders and both the exposure (DEX) and the outcome (in-hospital mortality) reaches 2.8 and 2.09, respectively, could the observed association be entirely explained away.

Patients with SIMI were divided into subgroups based on age, gender, SOFA score, CRRT use, SAPS II score, race, and potassium levels to evaluate the effect of DEX on 28-day mortality. The forest plot (Figure 3) illustrates the subgroup analysis findings, showcasing the impact of DEX on various patient groups. The results demonstrated that DEX significantly decreased 28-day mortality in certain subpopulations. For instance, patients aged ≤60 years (HR 0.365, 95% CI 0.214–0.622, p = 0.0002), male patients (HR 0.474, 95% CI 0.305–0.738, p = 0.0009), those with elevated SOFA scores (HR 0.399, 95% CI 0.261–0.610, p < 0.0001), patients not receiving CRRT (HR 0.681, 95% CI 0.495–0.937, p = 0.018), and those with higher SAPS II scores (HR 0.439, 95% CI 0.272–0.708, p = 0.0007) all exhibited notable protective effects. Additionally, significant benefits were identified in specific racial subgroups, such as race group 3 (HR 0.365, 95% CI 0.146–0.909, p = 0.030) and race group 4 (HR 0.485, 95% CI 0.268–0.876, p = 0.016). Patients with elevated potassium levels (HR 0.537, 95% CI 0.300–0.962, p = 0.037) also experienced reduced mortality risks. On the other hand, no significant protective effects were observed in patients aged >60 years (HR 0.915, 95% CI 0.616–1.360, p = 0.661), female patients (HR 0.926, 95% CI 0.595–1.441, p = 0.734), those with low SOFA scores (HR 1.080, 95% CI 0.671–1.739, p = 0.751), patients requiring CRRT (HR 0.191, 95% CI 0.040–0.917, p = 0.038), or patients with lower SAPS II scores (HR 0.773, 95% CI 0.513–1.167, p = 0.220). Similarly, patients in race group 0 (HR 1.641, 95% CI 0.103–26.258, p = 0.726) and race group 1 (HR 0.749, 95% CI 0.496– 1.132, p = 0.170), as well as those with normal or low potassium levels (HR 0.673, 95% CI 0.466–0.972, p = 0.035), did not demonstrate a significant reduction in mortality risk.In summary, this subgroup analysis highlights the potential protective effects of DEX in specific populations, underscoring the heterogeneity of the study cohort. These findings indicate that DEX may play an essential role in designing personalized treatment strategies for critically ill patients.

**Figure 3:**
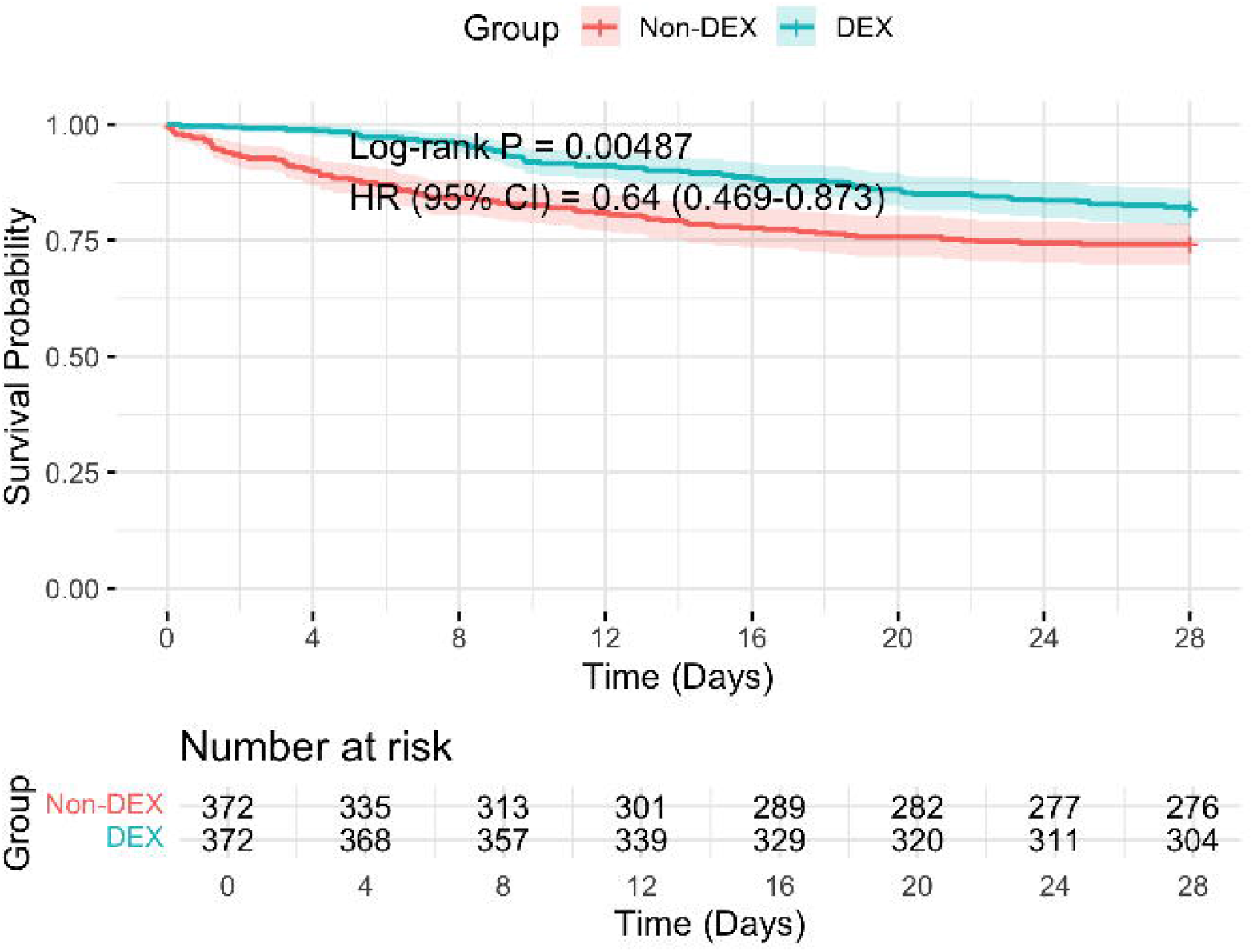
Forest plot illustrating the subgroup analysis of the association between DEX and 28-day mortality.

### External validation

A total of 1,282 patients were included in the external validation cohort. The baseline characteristics of these patients are shown in Table 4. We observed that the use of dexmedetomidine was associated with differences in in-hospital and 28-day mortality outcomes. Cox proportional hazards regression analysis indicated that dexmedetomidine use was potentially associated with reduced in-hospital and 28-day mortality.

For the 28-day mortality analysis, the unadjusted Cox regression model (Model 1) showed a hazard ratio (HR) of 0.49 (95% CI: 0.23–1.04, p = 0.061). After adjusting for age and gender (Model 2), the HR was 0.58 (95% CI: 0.27–1.22, p = 0.152). In the fully adjusted model, which accounted for age, gender, propofol, fentanyl, midazolam, and CRRT (Model 3), the HR was 0.49 (95% CI: 0.23–1.05, p = 0.068) (Table 5).

For in-hospital mortality, the unadjusted Cox regression model (Model 1) demonstrated that dexmedetomidine use significantly reduced risk, with an HR of 0.38 (95% CI: 0.17–0.86, p = 0.020). After adjusting for age and gender (Model 2), the HR was 0.44 (95% CI: 0.20–1.00, p = 0.049). In the fully adjusted model (Model 3), the HR was 0.45 (95% CI: 0.20–1.02, p = 0.055) (Table 5).

**Table 4:** Patient Demographics and Baseline Characteristics of the External Validation Cohort. Abbreviations: ICU: Intensive Care Unit; CRRT: Continuous Renal Replacement Therapy.

**Table 5:** Survival Outcomes of DEX Users and Non-Users in SIMI Patients from the External Validation Cohort. Abbreviations: HR: Hazard Ratio; CI: Confidence Interval.

## Discussion

In this retrospective cohort study, we assessed the impact and potential effects of DEX on 28-day hospital mortality and in-hospital mortality among patients with SIMI using propensity score matching (PSM) and Cox regression analysis. To enhance the accuracy and reliability of the findings, strict inclusion and exclusion criteria were applied. Patients with underlying cardiovascular conditions such as coronary artery disease or cardiomyopathy were excluded to minimize confounding contributions to myocardial injury. Similarly, individuals with pre-existing renal dysfunction were excluded to eliminate potential interference with troponin clearance. Additionally, only patients receiving DEX for 4 to 48 hours were included to reduce bias related to excessively short or prolonged drug administration durations, thereby ensuring the stability of the results.

The study results demonstrated that the use of DEX was associated with significantly lower 28-day hospital mortality and in-hospital mortality rates among ICU patients with SIMI, and our external validation cohort confirmed this finding. While DEX use was linked to prolonged ICU and overall hospital stays, this may reflect its role in improving hemodynamic stability and reducing mortality risk, ultimately allowing more time for treatment and recovery. Furthermore, no significant differences were observed between the DEX and non-DEX groups regarding the use of continuous renal replacement therapy (CRRT). To ensure the robustness of our findings, commonly used sedative and analgesic agents such as propofol, midazolam, and fentanyl were considered potential confounding factors. By employing propensity score matching, we effectively minimized the confounding effects of these medications on survival outcomes and length of stay.

Although previous studies have explored the mechanisms of DEX in SIMI large-scale cohort studies remain limited. This study, leveraging data from a large cohort, provides the first evidence supporting the potential value of DEX in reducing 28-day hospital mortality and in-hospital mortality. The findings underscore the significant role of DEX in improving the prognosis of patients with SIMI and highlight its importance as a therapeutic consideration in this critical condition.

The pathological characteristics of SIMI primarily include excessive infiltration of inflammatory cells, interstitial fibrosis, and mitochondrial damage^23^,Inflammation is a core feature of sepsis, and oxidative stress induced by intrinsic inflammatory responses can trigger lipid peroxidation, cause DNA damage, and exacerbate mitochondrial dysfunction, further worsening organ dysfunction and failure^24^. DEX, a highly selective α2-adrenergic receptor agonist, is widely used for its sedative, analgesic, anxiolytic, and anti-inflammatory properties in perioperative and intensive care settings^25^. In recent years, numerous studies have highlighted the organ-protective effects of DEX in sepsis, showing its potential to reduce mortality in patients with septic shock^26–28^. However, the specific mechanisms by which DEX influences sepsis-induced myocardial injury remain unclear.

Some studies suggest that the protective effects of DEX may be associated with its anti-inflammatory properties. Yimou et al.^29^ reported that early administration of DEX significantly reduced in-hospital mortality in patients with acute myocardial infarction, partly through the reduction of leukocyte-mediated inflammatory responses. Similarly, Tianyi et al. ^30^ demonstrated in a sepsis-induced rat model that DEX alleviates myocardial injury by promoting autophagy in cardiomyocytes, an effect that was significantly diminished when autophagy was inhibited. In contrast, other translational studies suggest that DEX may exert adverse effects under specific conditions. One study found that DEX administered after sepsis induction may directly act on α2A-adrenergic receptors in macrophages, cardiac fibroblasts, and cardiac endothelial cells, thereby exacerbating cardiac dysfunction and negatively impacting sepsis prognosis^31^ .Thus, while DEX demonstrates promising protective effects in SIMI, its precise mechanisms of action require further investigation to clarify the optimal timing, indications, and potential risks associated with its clinical use.

To explore the heterogeneity of treatment effects of DEX across different patient subgroups, further evaluate its potential protective effects in specific populations, and validate the robustness of our findings, we conducted a subgroup analysis. This analysis aimed to identify which patients with SIMI might benefit most from DEX treatment, thereby providing a scientific basis for the development of individualized treatment strategies.The results of the subgroup analysis revealed that DEX significantly reduced the 28-day in-hospital mortality risk in certain subgroups, including patients aged ≤60 years, males, those with high SOFA scores (≥8), patients not receiving CRRT, those with high SAPS II scores (≥50), and Black patients. Interestingly, patients benefited from DEX regardless of whether their serum potassium levels were high or low. These findings suggest that DEX may offer stronger protective effects in these specific subgroups.Sepsis is a life-threatening condition characterized by dysregulated host responses to infection, leading to organ dysfunction^18^. A previous study on human sepsis revealed that patients aged ≥70 years exhibited reduced gene expression in blood leukocytes related to cytokine signaling and innate and adaptive immunity compared to patients younger than 50 years^32^. Furthermore, elderly patients exhibit a blunted and easily dysregulated host response to infection, with significant differences in cytokine expression during sepsis compared to non-elderly patients^33^. Our study found that the use of DEX significantly reduced the 28-day in-hospital mortality risk for SIMI patients aged ≤60 years, suggesting that age may be a crucial factor influencing the therapeutic efficacy of DEX. This may be attributed to younger patients possessing stronger immune responses and physiological reserves compared to older patients, allowing them to derive greater benefits from DEX’s protective effects against sepsis-induced myocardial injury. Moreover, younger patients likely have more responsive immune regulatory mechanisms, enabling a more effective reaction to the anti-inflammatory and sedative properties of DEX, thereby mitigating inflammation and oxidative stress-induced damage.Additionally, DEX demonstrated significant protective effects for male SIMI patients.

Currently, research on the relationship between sepsis mortality and gender remains inconsistent on the international stage^34–39^. Most of the evidence suggests that the protective effects of DEX are mediated through its ability to reduce excessive inflammation by lowering the expression of pro-inflammatory cytokines such as interleukin (IL)-6 and tumor necrosis factor (TNF)-α^40^. While male patients typically exhibit elevated levels of these pro-inflammatory cytokines, female patients tend to have higher levels of the anti-inflammatory cytokine IL-10, which is considered a predictive marker of sepsis severity ^41,42^. This immunoregulatory difference may make male patients more susceptible to inflammatory states. DEX, with its well-documented anti-inflammatory properties, may provide significant protective benefits to male patients by mitigating the excessive release of pro-inflammatory cytokines. This could explain the reduced mortality risk observed in male patients with SIMI, highlighting the potential for DEX to serve as a gender-specific therapeutic strategy.Patients with high Simplified Acute Physiology Score II (SAPS II ≥50) and Sequential Organ Failure Assessment (SOFA ≥8) are generally representative of the most critically ill populations, with SIMI often accompanied by multi-organ dysfunction and a higher risk of mortality. The SOFA score is a widely used tool to assess organ dysfunction in sepsis and is strongly correlated with multi-organ failure and mortality rates^43^ . Similarly, the SAPS II score integrates various physiological and laboratory parameters, providing a comprehensive measure of illness severity and a reliable predictor of mortality, especially among high-risk populations^44^ DEX, as a highly selective α2-adrenergic receptor agonist, exerts organ-protective effects through mechanisms such as suppressing sympathetic activity, modulating G-protein and intracellular signaling pathways, and inhibiting adenylate cyclase activity^45^ .These mechanisms collectively improve myocardial function, reduce oxidative stress and inflammation, and ultimately lower mortality risks in SIMI patients.Our study also revealed that Black patients demonstrated significant survival benefits from DEX treatment during subgroup analysis. This observation may be linked to specific physiological and biochemical differences. Research has shown that pain perception varies across racial groups, with African American patients generally reporting higher pain scores than White patients^46^ . This heightened pain perception could reflect an increased need for sedative and anti-inflammatory medications, potentially explaining why Black patients derive greater benefit from DEX treatment.Although DEX exhibits high protein binding, limiting its clearance by continuous renal replacement therapy (CRRT), studies suggest that certain CRRT circuits may adsorb drugs and affect their effective concentration. For instance, the concentrations of ketamine and DEX have been shown to be significantly impacted in extracorporeal life support (ECLS) circuits^47^. This implies that the pharmacological efficacy of DEX may be diminished in patients receiving CRRT, underscoring the need for careful consideration of its use in such settings.

This study has several limitations that need to be acknowledged. First, we utilized the MIMIC-IV database, which, although rich in critical care patient data, represents a specific ICU population in the United States. This may limit the generalizability of our findings to other regions and healthcare systems. Additionally, external validation was not performed in this study, and future research should include cohorts from Asian populations and conduct multi-center trials to confirm the broader applicability of the results. Second, the study only included patients who received DEX for 4 to 48 hours, which may not fully reflect the efficacy of the drug under different dosages and durations of administration, potentially affecting the generalizability of the results. Third, although we reduced baseline differences between groups using propensity score matching (PSM), the matched sample size was relatively small, which may have limited the statistical power. Lastly, as a retrospective study, residual confounding factors cannot be completely excluded. Future large-scale prospective studies are needed to further validate our findings and explore the optimal use of DEX in patients with SIMI.

## Conclusion

This study found that DEX may reduce the 28-day in-hospital mortality and overall in-hospital mortality in patients with sepsis-induced myocardial injury, providing preliminary evidence for its clinical application. These findings may assist healthcare professionals in deciding whether to use DEX, but further randomized controlled trials are needed to confirm these results.

## Data Availability

The data used in this study were obtained from the publicly accessible MIMIC-IV database (https://mimic.mit.edu/docs/iv/), with all data anonymized to protect patient privacy.Research data are available in accordance with the database access policy.

## Abbreviations

SIMI: sepsis-induced myocardial injury
DEX: dexmedetomidine
ICU: intensive care unit
SOFA: Sequential Organ Failure Assessment
SAPS II: Simplified Acute Physiology Score II
PSM: propensity score matching
HR: hazard ratio
CI: confidence interval
CRRT: continuous renal replacement therapy
ECLS: extracorporeal life support system

## Ethical Approval and Consent to Participate

In compliance with the Health Insurance Portability and Accountability Act (HIPAA), all patient information in the MIMIC-IV database has been anonymized to protect privacy. Therefore, no additional patient consent or ethical approval was required for this study. Zhang Xinyi, a member of the research team, obtained access to the database and was responsible for data extraction (Authorization Code: 10320923).

## Author Contributions

Zhang Xinyi, Guo Xiaoyan, and Zhao Zhongxing were responsible for data analysis and manuscript writing. Li Wei, Chen Huaiyu, and Li Hongyan provided statistical support and methodological guidance. Lin Jiandong and Lin Mingrui supervised the study design and critically reviewed the manuscript. All authors read and approved the final manuscript.

