## Supplemental Table 1 for "Impact of Dexmedetomidine on the Prognosis of Patients with Sepsis-Induced Myocardial Injury: A Retrospective Cohort Study"

TABLE 1：Baseline Characteristics Between DEX and Non-DEX Groups Before and After Propensity Score Matching

| Characteristics | Non-DEX group（n=372） | DEX group（n=372） | SMD | p | Non-DEX group（n=3546） | DEX group（n=375） | SMD | p |
| --- | --- | --- | --- | --- | --- | --- | --- | --- |
| Demographics |  |  |  |  |  |  |  |  |
| Race (%) |  |  | 0.179 | 0.116 |  |  | 0.26 | <0.001 |
| Asian | 12 ( 3.2) | 7 ( 1.9) |  |  | 126 ( 3.6) | 7 ( 1.9) |  |  |
| White | 207 ( 55.6) | 219 ( 58.9) |  |  | 2168 ( 61.1) | 219 ( 58.4) |  |  |
| Black | 85 ( 22.8) | 64 ( 17.2) |  |  | 774 ( 21.8) | 65 ( 17.3) |  |  |
| Unknown | 68 ( 18.3) | 82 ( 22.0) |  |  | 478 ( 13.5) | 84 ( 22.4) |  |  |
| Gender (%) |  |  | 0.1 | 0.2 |  |  | 0.166 | 0.003 |
| Male | 221 ( 59.4) | 239 ( 64.2) |  |  | 2001 ( 56.4) | 242 ( 64.5) |  |  |
| Female | 151 ( 40.6) | 133 ( 35.8) |  |  | 1545 ( 43.6) | 133 ( 35.5) |  |  |
| Age (years) | 60.00 [49.75, 71.00] | 61.00 [47.75, 70.25] | 0.003 | 0.972 | 63.00 [51.25, 75.75] | 61.00 [47.50, 70.00] | 0.221 | <0.001 |
| Weight (kg) | 80.28 [69.00, 95.63] | 80.45 [70.00, 95.91] | 0.084 | 0.643 | 77.45 [65.00, 92.18] | 80.45 [70.00, 95.90] | 0.171 | <0.001 |
| Vital Signs |  |  |  |  |  |  |  |  |
| Heart rate (mean, bpm) | 86.19 [75.08, 99.45] | 87.08 [74.94, 99.07] | 0.015 | 0.824 | 88.88 [76.47, 101.73] | 87.07 [74.91, 99.18] | 0.095 | 0.071 |
| SBP (mean, mmHg) | 113.17 [105.85, 125.94] | 114.28 [105.05, 127.20] | 0.064 | 0.433 | 113.47 [104.89, 126.74] | 114.25 [104.90, 127.05] | 0.063 | 0.299 |
| DBP (mean, mmHg) | 62.06 [56.25, 68.22] | 62.03 [57.77, 69.62] | 0.084 | 0.177 | 62.43 [56.20, 69.65] | 62.00 [57.78, 69.67] | 0.014 | 0.521 |
| MBP (mean, mmHg) | 76.76 [72.30, 85.13] | 78.50 [72.12, 85.11] | 0.059 | 0.342 | 76.88 [70.89, 85.08] | 78.48 [72.14, 85.10] | 0.086 | 0.058 |
| Respiratory rate (mean, bpm) | 19.90 [17.47, 23.58] | 20.04 [17.38, 23.30] | 0.05 | 0.609 | 19.78 [17.10, 23.25] | 20.07 [17.38, 23.30] | 0.01 | 0.625 |
| Temperature (mean, °C) | 37.11 [36.76, 37.52] | 37.10 [36.75, 37.53] | 0.005 | 0.918 | 36.91 [36.60, 37.31] | 37.13 [36.75, 37.56] | 0.37 | <0.001 |
| SpO₂ (mean, %) | 97.92 [96.25, 99.04] | 97.67 [96.23, 98.88] | 0.061 | 0.222 | 97.25 [95.77, 98.61] | 97.67 [96.23, 98.90] | 0.222 | <0.001 |
| Severity Scores |  |  |  |  |  |  |  |  |
| SOFA | 6.00 [4.00, 10.00] | 6.00 [4.00, 9.25] | 0.015 | 0.863 | 5.00 [3.00, 9.00] | 6.00 [4.00, 9.50] | 0.132 | 0.003 |
| SAPS II | 38.00 [31.00, 49.25] | 39.00 [30.00, 51.00] | 0.05 | 0.652 | 40.00 [31.00, 51.00] | 39.00 [30.00, 50.50] | 0.105 | 0.162 |
| Laboratory Tests |  |  |  |  |  |  |  |  |
| Hematocrit (% min) | 30.90 [26.48, 35.20] | 31.20 [26.28, 35.62] | 0.015 | 0.897 | 30.60 [25.30, 35.40] | 31.40 [26.20, 35.70] | 0.053 | 0.294 |
| Platelets (×10⁹/L min) | 160.00 [105.75, 227.25] | 171.50 [102.75, 233.25] | 0.008 | 0.512 | 168.00 [107.00, 236.00] | 172.00 [103.00, 237.00] | 0.02 | 0.841 |
| WBC (×10⁹/L min) | 9.60 [6.47, 12.80] | 9.80 [6.40, 13.12] | 0.022 | 0.948 | 9.80 [6.70, 13.80] | 9.80 [6.40, 13.10] | 0.08 | 0.253 |
| WBC (×10⁹/L max) | 14.00 [10.07, 18.33] | 13.75 [10.17, 18.90] | 0.051 | 0.611 | 14.00 [9.80, 19.30] | 13.60 [10.10, 18.90] | 0.054 | 0.718 |
| Blood urea nitrogen (BUN, mg/dL) | 19.00 [13.00, 28.00] | 19.50 [14.00, 29.00] | 0.006 | 0.575 | 22.00 [15.00, 35.00] | 19.00 [13.00, 29.00] | 0.224 | <0.001 |
| Creatinine (mg/dL max) | 1.00 [0.80, 1.50] | 1.00 [0.80, 1.50] | 0.044 | 0.603 | 1.10 [0.80, 1.60] | 1.00 [0.80, 1.50] | 0.075 | 0.005 |
| Calcium (mg/dL min) | 7.80 [7.30, 8.30] | 7.90 [7.40, 8.30] | 0.016 | 0.362 | 7.90 [7.30, 8.40] | 7.90 [7.40, 8.30] | 0.049 | 0.567 |
| Calcium (mg/dL max) | 8.40 [7.90, 8.90] | 8.50 [8.00, 9.00] | 0.018 | 0.288 | 8.50 [8.00, 9.00] | 8.50 [8.00, 9.00] | 0.067 | 0.673 |
| Sodium (mmol/L min) | 137.00 [134.00, 140.00] | 137.00 [134.00, 140.00] | 0.023 | 0.716 | 137.00 [134.00, 140.00] | 137.00 [134.00, 140.00] | 0.049 | 0.845 |
| Sodium (mmol/L max) | 141.00 [138.00, 143.00] | 140.00 [138.00, 143.00] | 0.016 | 0.809 | 140.00 [137.00, 143.00] | 141.00 [138.00, 143.00] | 0.066 | 0.735 |
| Potassium (mmol/L min) | 3.70 [3.40, 4.00] | 3.70 [3.40, 4.00] | 0.037 | 0.789 | 3.80 [3.40, 4.10] | 3.70 [3.40, 4.00] | 0.068 | 0.219 |
| Potassium (mmol/L max) | 4.40 [4.00, 4.90] | 4.40 [4.00, 4.90] | 0.024 | 0.778 | 4.40 [4.00, 4.90] | 4.40 [4.00, 4.90] | 0.031 | 0.742 |
| Glucose (mg/dL min) | 113.00 [94.00, 135.25] | 113.00 [94.00, 136.25] | 0.078 | 0.879 | 113.00 [94.00, 137.00] | 113.00 [94.00, 136.00] | 0.028 | 0.838 |
| Glucose (mg/dL max) | 150.00 [126.75, 195.25] | 153.00 [123.75, 200.50] | 0.033 | 0.887 | 156.00 [126.00, 211.00] | 153.00 [123.00, 200.00] | 0.126 | 0.177 |
| Glucose (mg/dL mean) | 131.53 [112.33, 157.75] | 132.33 [112.04, 161.48] |  |  | 135.02 [112.33, 168.71] | 132.01 [111.69, 161.25] | 0.025 | 0.05 |
| Interventions |  |  |  |  |  |  |  |  |
| fentanyl(%) |  |  | <0.001 | 1 |  |  | 0.948 | <0.001 |
| No | 56 ( 15.1) | 56 ( 15.1) |  |  | 1982 ( 55.9) | 56 ( 14.9) |  |  |
| Yes | 316 ( 84.9) | 316 ( 84.9) |  |  | 1564 ( 44.1) | 319 ( 85.1) |  |  |
| midazolam(%) |  |  | 0.038 | 0.66 |  |  | 0.475 | <0.001 |
| No | 195 ( 52.4) | 188 ( 50.5) |  |  | 2584 ( 72.9) | 189 ( 50.4) |  |  |
| Yes | 177 ( 47.6) | 184 ( 49.5) |  |  | 962 ( 27.1) | 186 ( 49.6) |  |  |
| propofol(%) |  |  | 0.065 | 0.448 |  |  | 1.177 | <0.001 |
| No | 341 ( 91.7) | 334 ( 89.8) |  |  | 2065 ( 58.2) | 38 ( 10.1) |  |  |
| Yes | 360 ( 96.8) | 363 ( 97.6) |  |  | 1481 ( 41.8) | 337 ( 89.9) |  |  |
| CRRT (%) |  |  |  |  |  |  | 0.036 | 0.63 |
| No | 360 ( 96.8) | 363 ( 97.6) | 0.049 | 0.658 | 3440 ( 97.0) | 366 ( 97.6) |  |  |
| Yes | 12 ( 3.2) | 9 ( 2.4) |  |  | 106 ( 3.0) | 9 ( 2.4) |  |  |

Abbreviations: SMD: Standardized Mean Difference; SBP: Systolic Blood Pressure; DBP: Diastolic Blood Pressure; MBP: Mean Blood Pressure; SpO₂: Oxygen Saturation; SOFA: Sequential Organ Failure Assessment; SAPSII: Simplified Acute Physiology Score II; WBC: White Blood Cell; BUN: Blood Urea Nitrogen; CRRT: Continuous Renal Replacement Therapy.
