## Supplemental Table 2 for "Impact of Dexmedetomidine on the Prognosis of Patients with Sepsis-Induced Myocardial Injury: A Retrospective Cohort Study"

TABLE 2：Comparison of Clinical Outcomes Between DEX and Non-DEX Groups Before and After Propensity Score Matching

| Outcomes | Non-DEX group（n=372） | DEX group（n=372） | SMD | p | Non-DEX group（n=3546） | DEX group（n=375） | SMD | p |
| --- | --- | --- | --- | --- | --- | --- | --- | --- |
| In-hospital mortality (%) |  |  | 0.292 | <0.001 |  |  | 0.295 | <0.001 |
| No | 271 ( 72.8) | 315 ( 84.7) |  |  | 2584 ( 72.9) | 318 ( 84.8) |  |  |
| Yes | 101 ( 27.2) | 57 ( 15.3) |  |  | 962 ( 27.1) | 57 ( 15.2) |  |  |
| 28-day mortality (%) |  |  | 0.292 | <0.001 |  |  | 0.249 | <0.001 |
| No | 276 ( 74.2) | 304 ( 81.7) |  |  | 2532 ( 71.4) | 307 ( 81.9) |  |  |
| Yes | 96 ( 25.8) | 68 ( 18.3) |  |  | 1014 ( 28.6) | 68 ( 18.1) |  |  |
| ICU length of stay (hours) | 139.67 [65.31, 280.00] | 191.28 [112.55, 321.98] | 0.23 | <0.001 | 89.51 [45.61, 192.84] | 190.27 [112.29, 320.66] | 0.47 | <0.001 |
| Hospital length of stay (hours) | 311.42 [142.11, 525.53] | 385.45 [238.86, 613.40] | 0.244 | <0.001 | 227.97 [116.13, 445.62] | 384.37 [237.72, 612.34] | 0.338 | <0.001 |

Abbreviations:  ICU: Intensive Care Unit；p: p-value (statistical significance)
