## Supplemental Table 3 for "Impact of Dexmedetomidine on the Prognosis of Patients with Sepsis-Induced Myocardial Injury: A Retrospective Cohort Study"

**Table 3: Survival Outcomes of DEX Users and Non-Users in SIMI Patients Before and After Propensity Score Matching**

| **Categories** | **28-day hospital mortality** | **In-hospital mortality** |
| --- | --- | --- |
| **Before PSM** | HR (95% CI,p-value) | |
| Model 1 | 0.57 (0.447–0.731, p = 8.01e-06) | 0.42 (0.318–0.544, p = 1.33e-10) |
| Model 2 | 0.53 (0.411–0.674, p = 3.53e-07) | 0.40 (0.309–0.529, p = 3.81e-11) |
| Model 3 | 0.57 (0.443–0.739, p = 1.94e-05) | 0.44 (0.333–0.579, p = 5.59e-09) |
| **After PSM** | HR (95% CI,p-value) | |
| Model 1 | 0.64 (0.469–0.873, p = 0.00487) | 0.46 (0.329–0.632, p = 2.29e-06) |
| Model 2 | 0.65 (0.477–0.889, p = 0.00697) | 0.48 (0.345–0.663, p = 9.81e-06) |
| Model 3 | 0.58 (0.417–0.798, p = 0.00089) | 0.41 (0.288–0.577, p = 4.13e-07) |

Abbreviations:  HR: Hazard Ratio；CI: Confidence Interval ；p-value: Probability Value

Model 1: Unadjusted model, including only the DEX and non-DEX group variables.

Model 2: Adjusted for key demographic variables, including age, gender, and SOFA、SAPSII.

Model 3: Fully adjusted model, accounting for confounding variables with p-value <0.05 in univariate analysis. These variables include age, gender, ethnicity, CRRT, propofol, midazolam, fentanyl, respiratory rate, SpO₂, BUN, creatinine, SOFA, and others.
