## Supplemental Table 4 for "Impact of Dexmedetomidine on the Prognosis of Patients with Sepsis-Induced Myocardial Injury: A Retrospective Cohort Study"

Table 4 ：Patient Demographics and Baseline Characteristics of the External Validation Cohort

| Characteristic | dex group | | p-value |
| --- | --- | --- | --- |
|  | 0, N = 1,247^1^ | 1, N = 35^1^ |  |
| Age(years) | 69 (59, 80) | 59 (49, 64) | <0.001 |
| Gender (%) |  |  | 0.938 |
| Male | 740 (59.3%) | 21 (60.0%) |  |
| Female | 507 (40.7%) | 14 (40.0%) |  |
| Fentanyl (%) |  |  | <0.001 |
| No | 604 (48.4%) | 2 (5.7%) |  |
| Yes | 643 (51.6%) | 33 (94.3%) |  |
| Midazolam (%) |  |  | <0.001 |
| No | 694 (55.7%) | 7 (20.0%) |  |
| Yes | 553 (44.3%) | 28 (80.0%) |  |
| Propofol (%) |  |  | <0.001 |
| No | 871 (69.8%) | 6 (17.1%) |  |
| Yes | 376 (30.2%) | 29 (82.9%) |  |
| Crrt(%) |  |  | 0.155 |
| No | 1,124 (90.1%) | 29 (82.9%) |  |
| Yes | 123 (9.9%) | 6 (17.1%) |  |
| ICU length of stay (hours) | 5 (2, 11) | 12 (8, 18) | <0.001 |
| Hospital length of stay (hours) | 11 (5, 20) | 15 (12, 22) | 0.001 |
| In-hospital mortality (%) |  |  | 0.020 |
| No | 794 (63.7%) | 29 (82.9%) |  |
| Yes | 453 (36.3%) | 6 (17.1%) |  |
| 28-day mortality (%) |  |  | 0.053 |
| 0 | 800 (64.2%) | 28 (80.0%) |  |
| 1 | 447 (35.8%) | 7 (20.0%) |  |

Abbreviations: ICU: Intensive Care Unit; CRRT: Continuous Renal Replacement Therapy;
