## Supplemental Table 5 for "Impact of Dexmedetomidine on the Prognosis of Patients with Sepsis-Induced Myocardial Injury: A Retrospective Cohort Study"

**Table 5：Survival Outcomes of DEX Users and Non-Users in SIMI Patients from the Validation Cohort**

| Outcome | Model | HR (95% CI) | P-value |
| --- | --- | --- | --- |
| 28-Day Mortality | Model 1 | 0.49 (0.23–1.04) | 0.061 |
|  | Model 2 | 0.58 (0.27–1.22) | 0.152 |
|  | Model 3 | 0.49 (0.23–1.05) | 0.068 |
| In-Hospital Mortality | Model 1 | 0.38 (0.17–0.86) | 0.02 |
|  | Model 2 | 0.44 (0.20–1.00) | 0.049 |
|  | Model 3 | 0.45 (0.20–1.02) | 0.055 |

**Abbreviations:** HR: Hazard Ratio; CI: Confidence Interval.
